# Structured Patterns of Muscle Involvement in CAV3-Related Myopathy Revealed by Whole-Body CT Imaging

**DOI:** 10.64898/2026.06.03.26354504

**Authors:** Francia Victoria A. De Los Reyes, Shinichiro Hayashi, Yoshihiko Saito, Megumu Ogawa, Yasushi Oya, Satoru Noguchi, Ichizo Nishino

**Affiliations:** Department of Neuromuscular Research, National Institute of Neuroscience, National Center of Neurology and Psychiatry, Kodaira, Tokyo, Japan; Department of Genome Medicine Development, Medical Genome Center, National Center of Neurology and Psychiatry, Tokyo, Japan; Department of Neurology, National Center Hospital, National Center of Neurology and Psychiatry, Tokyo, Japan

**Keywords:** caveolinopathy, CAV3 mutation, skeletal muscle CT, modified Mercuri score, multivariate modeling

## Abstract

Caveolinopathies caused by CAV3 mutations present with heterogeneous clinical phenotypes ranging from asymptomatic hyperCKemia to limb-girdle–type muscular dystrophy. Although prior imaging studies have described commonly affected muscles, structured modeling of muscle involvement patterns in caveolinopathy has not been established. We analyzed whole-body skeletal muscle computed tomography imaging in eight patients with pathogenic or likely pathogenic CAV3 variants, comprising 14 imaging study samples. Fat infiltration across 43 muscles was graded using modified Mercuri scores. Computational multivariate analysis—including principal component analysis, clustering, and pseudotime modeling—was applied to characterize severity staging and distribution patterns.

A statistically supported, stage-dependent continuum of muscle involvement was identified. Most samples demonstrated a distributed limb-girdle–predominant pattern with coordinated progression across muscle clusters. In contrast, one patient (three samples in longitudinal series) exhibited a compartment-restricted thigh-dominant pattern characterized by early posterior and medial thigh involvement. Rectus femoris showed consistent stage-dependent progression, while greater medial gastrocnemius involvement was associated with advanced severity. None of the patients exhibited clinical evidence of rippling muscle disease.

These findings suggest that integrating semi-quantitative imaging with computational modeling may provide an objective framework for characterizing muscle involvement patterns in CAV3-related myopathy.

**Highlights:**

- This is a whole-body muscle imaging analysis of caveolinopathy patients.
- Multivariate modeling was applied to CT-based modified Mercuri scores.
- Rectus femoris involvement may be useful as a progression indicator.
- Two distinct muscle involvement patterns were described.
- Greater medial gastrocnemius involvement may reflect more advanced disease.

## 1. Introduction

Caveolinopathies constitute a group of neuromuscular disorders characterized by a relatively heterogeneous yet distinct clinical phenotype, primarily attributed to mutations in the *CAV3* gene localized to chromosome 3p25.^1^ The diversity of genetic variations in *CAV3* has been linked to a spectrum of clinical manifestations, including asymptomatic elevated creatine kinase (CK), post-exertional myalgia, rippling muscle disease, limb-girdle type muscular dystrophy, and distal myopathy.^2–4^ Information on the genotype-phenotype correlation may be limited due to the heterogeneity of clinical presentation; some patients with the same variant in the same family may present with different phenotypes. ^3^

The symptomatology and phenotypic variability observed among affected individuals may be attributable to the domain of the caveolin-3 protein affected by the variant because involvement of certain regions of the domain may differentially disrupt oligomerization and interaction with other membrane proteins, thus affecting sarcolemmal integrity and altering the muscle’s adaptive response to repetitive mechanical stress.^5,6^ More specifically, its interaction with the dystrophin-associated glycoprotein complex enables the maintenance of muscle fiber structure by linking the extracellular matrix with the cytoskeleton. Thus, the underlying pathology that has been associated with derangements in cellular mechanisms due to the perturbation of cytoskeletal anchorage to the extracellular matrix and failure of modulation of intracellular signaling pathways may be responsible for progressive myofiber degeneration and fat replacement observable in skeletal muscle imaging studies.^7^

Prior studies have described muscle involvement patterns in individual patients or specific variants; however, it remains unclear whether a shared progression architecture exists across phenotypically heterogeneous CAV3-related myopathy. Because prior reports have relied on descriptive, patient-specific observations rather than systematic whole-body computational analysis, the identification of consistent involvement patterns and clinically informative muscle markers across the disease spectrum has not been established.^8,9^

The present study addresses this gap by applying whole-body skeletal muscle CT imaging combined with multivariate computational modeling across a cohort with heterogeneous CAV3 variants and clinical presentations. Specifically, this investigation aims to determine: (1) whether individuals with *CAV3* variants demonstrate a consistent pattern of muscle involvement and disease progression despite differences in initial clinical symptoms; (2) whether specific muscle groups are preferentially affected and may serve as early indicators of disease activity or progression markers; and (3) whether particular *CAV3* variants, especially those affecting defined protein domains, correlate with selective muscle involvement.

## 2. Materials and Methods

### 2.1 Subjects

A total of eight patients were included in this cohort, representing all individuals with a pathogenic or likely pathogenic CAV3 variant evaluated at the National Center of Neurology and Psychiatry (NCNP)—a nationwide referral center for neuromuscular diseases in Japan—for whom blood samples and skeletal muscle imaging studies were available during the study ascertainment period (January 1, 2001, to December 31, 2024). The NCNP neuromuscular repository was established in 1978; however, the present study cohort was restricted to cases with complete imaging and biospecimen availability within 2001–2024. All CAV3 variants had been previously identified for diagnostic purposes.

Two patients underwent serial imaging studies at intervals of ≥ 1 year. Each imaging study separated by one year or more was treated as an independent analytical time point and assigned a unique identifier to enable assessment of disease progression.

This study was approved by the Ethical Committee of the NCNP, and written informed consent for diagnostic and research use of clinical data was obtained from all participants.

### 2.2 Epidemiological Data

Demographic data, clinical history, physical examination findings, muscle pathology, and genetic mutation status were retrieved from archived paper and digital records maintained in the NCNP muscle repository. All patient-identifying information was excluded.

Variables analyzed included age at initial consultation, sex, duration of symptoms prior to consultation, initial clinical presentation, serum CK levels, and the specific CAV3 variant identified. These data are summarized in Table 1.

**Table 1.**
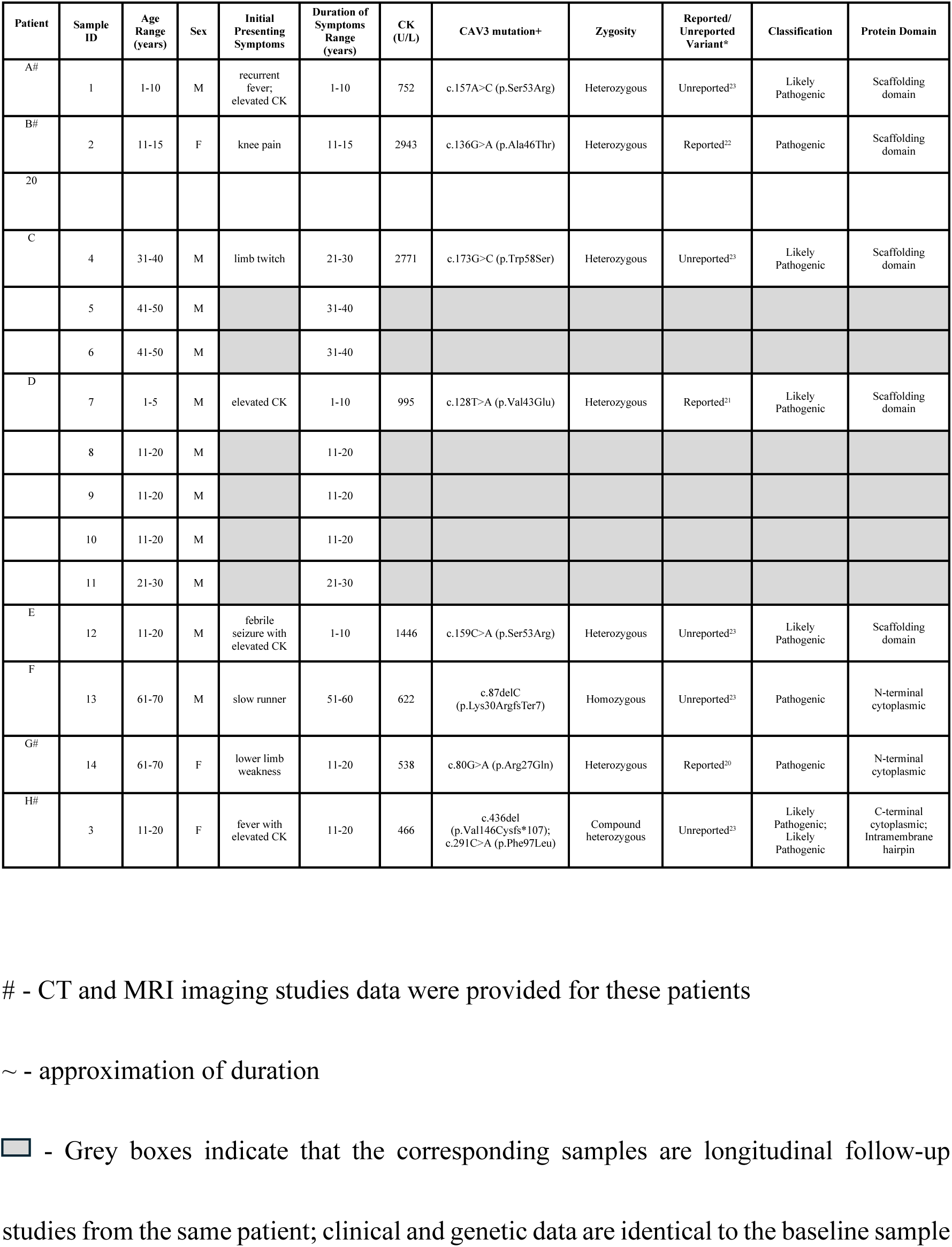
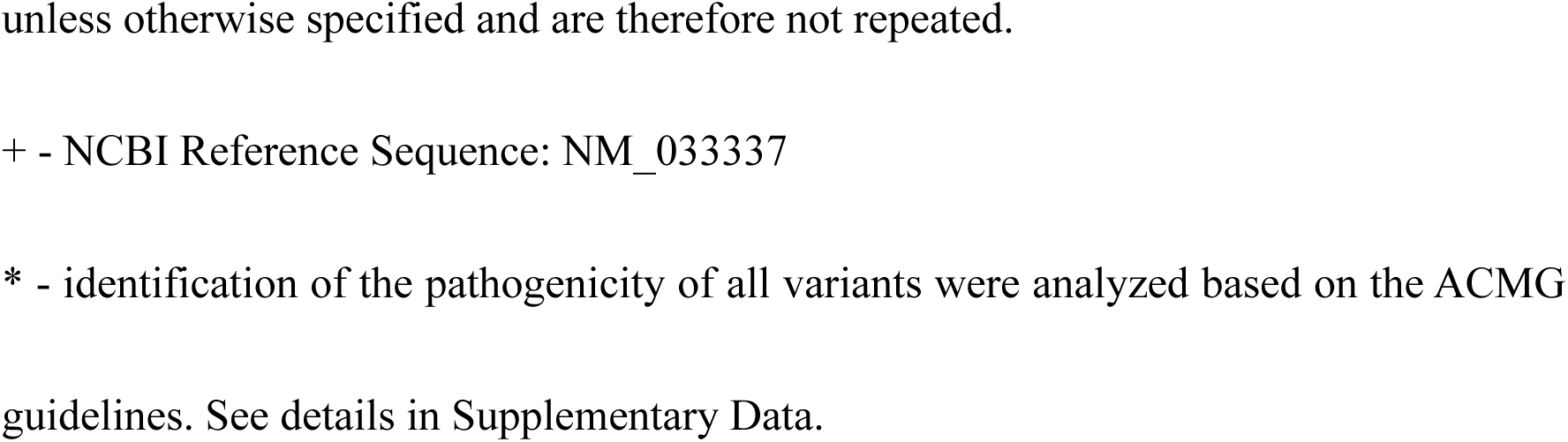
Demographics and Mutation Information of Patients.

| Patient | Sample ID | Age Range (years) | Sex | Initial Presenting Symptoms | Duration of Symptoms Range (years) | CK (U/L) | CAV3 mutation<sup>+</sup> | Zygosity | Reported/ Unreported Variant<sup>*</sup> | Classification | Protein Domain |
| --- | --- | --- | --- | --- | --- | --- | --- | --- | --- | --- | --- |
| A# | 1 | 1-10 | M | recurrent fever; elevated CK | 1-10 | 752 | c.157A>C (p.Ser53Arg) | Heterozygous | Unreported<sup>23</sup> | Likely Pathogenic | Scaffolding domain |
| B# | 2 | 11-15 | F | knee pain | 11-15 | 2943 | c.136G>A (p.Ala46Thr) | Heterozygous | Reported<sup>22</sup> | Pathogenic | Scaffolding domain |
| 20 |  |  |  |  |  |  |  |  |  |  |  |
| C | 4 | 31-40 | M | limb twitch | 21-30 | 2771 | c.173G>C (p.Trp58Ser) | Heterozygous | Unreported<sup>23</sup> | Likely Pathogenic | Scaffolding domain |
|  | 5 | 41-50 | M |  | 31-40 |  |  |  |  |  |  |
|  | 6 | 41-50 | M |  | 31-40 |  |  |  |  |  |  |
| D | 7 | 1-5 | M | elevated CK | 1-10 | 995 | c.128T>A (p.Val43Glu) | Heterozygous | Reported<sup>21</sup> | Likely Pathogenic | Scaffolding domain |
|  | 8 | 11-20 | M |  | 11-20 |  |  |  |  |  |  |
|  | 9 | 11-20 | M |  | 11-20 |  |  |  |  |  |  |
|  | 10 | 11-20 | M |  | 11-20 |  |  |  |  |  |  |
|  | 11 | 21-30 | M |  | 21-30 |  |  |  |  |  |  |
| E | 12 | 11-20 | M | febrile seizure with elevated CK | 1-10 | 1446 | c.159C>A (p.Ser53Arg) | Heterozygous | Unreported<sup>23</sup> | Likely Pathogenic | Scaffolding domain |
| F | 13 | 61-70 | M | slow runner | 51-60 | 622 | c.87delC (p.Lys30ArgfsTer7) | Homozygous | Unreported<sup>23</sup> | Pathogenic | N-terminal cytoplasmic |
| G# | 14 | 61-70 | F | lower limb weakness | 11-20 | 538 | c.80G>A (p.Arg27Gln) | Heterozygous | Reported<sup>20</sup> | Pathogenic | N-terminal cytoplasmic |
| H# | 3 | 11-20 | F | fever with elevated CK | 11-20 | 466 | c.436del (p.Val146Cysfs*107); c.291C>A (p.Phe97Leu) | Compound heterozygous | Unreported<sup>23</sup> | Likely Pathogenic; Likely Pathogenic | C-terminal cytoplasmic; Intramembrane hairpin |

### 2.3 Muscle Imaging

Computed tomography (CT) constituted the primary imaging modality in clinical practice. Although magnetic resonance imaging (MRI) was available for a subset of patients, all individuals underwent CT. Semi-quantitative analysis was therefore performed exclusively on CT images to ensure methodological uniformity. CT acquisition parameters were as follows: slice thickness 3–10 mm; matrix size 512 × 512; in-plane pixel spacing 0.390 × 0.390 mm to 1.269 × 1.269 mm.

Whole-body skeletal muscle imaging analysis was defined as evaluation of axial CT images encompassing 43 muscles across the neck, upper limb, trunk, pelvis, thigh, and lower leg. Cardiac imaging was not included.

Muscle identification followed predefined osseous landmarks^10–12^ applied consistently across all scans:

- Neck: mid-cervical vertebral levels
- Shoulder/upper trunk: humeral head and scapular spine
- Thoracic trunk: mid-thoracic vertebral bodies
- Abdominal wall: third lumbar vertebra (L3)
- Pelvis: acetabular roof and femoral head
- Thigh: region between the inferior ischial tuberosity and distal femoral condyles (ischial tuberosity defining pelvic–thigh boundary)
- Lower leg: proximal tibial plateau and fibular head, with mid-calf sections for gastrocnemius and soleus

For each muscle, the axial slice demonstrating the largest cross-sectional area and greatest degree of fatty infiltration was selected for scoring.

The muscles assessed were as follows:

- Neck muscles – cervical paraspinal (CPs), sternocleidomastoid (Scm), levator scapulae (LS), splenius capitis (SC)
- Upper arm – biceps brachii (BB), triceps brachii (TB), brachialis (Br)
- Shoulder and upper chest wall / trunk – teres major (TM), pectoralis major (PM), deltoid (Del), infraspinatus (Infs), subscapularis (Subs), trapezius (Tpz), rhomboid (Rh), serratus anterior (SA), thoracic paraspinal (TPs)
- Abdominal wall – iliocostalis lumborum (IcL), longissimus (Lo), multifidus (Mf), and rectus abdominis (RA)
- Pelvic muscles – gluteus maximus (GMx), gluteus medius (GMd), and gluteus minimus (GMn)
- Thigh muscles – rectus femoris (RF), sartorius (Sar), gracilis (Gr), vastus intermedius (VI), vastus lateralis (VL), vastus medialis (VM), adductor longus (AL), adductor magnus (AM), semimembranosus (Sm), semitendinosus (St), long head of the biceps femoris (BFL), and short head of the biceps femoris (BFS)
- Lower leg muscles – tibialis anterior (TA), tibialis posterior (TP), extensor digitorum longus (EDL), fibularis longus (FL), fibularis brevis (FB), soleus (Sol), medial gastrocnemius (GcM) and lateral gastrocnemius (GcL).

Fat infiltration in each muscle was assessed using a modified version of the Mercuri score (mMS). Prior work supports comparable interpretation of mMS fat replacement grading across CT and MRI.^13,14^

The mMS grading system was defined as follows:

- **0** – Approximating normal muscle appearance
- **1** – Mildly decreased attenuation compared with normal muscle
- **2** – Moderately decreased attenuation involving <50% of the muscle
- **3** – Severely decreased attenuation involving >50% of the muscle
- **4** – Complete fatty replacement or marked low-attenuation change

Efforts were made to evaluate all target muscles using the available CT imaging data. Muscles lacking sufficient imaging coverage were noted and included in descriptive and hierarchical analyses of mMS scores but were excluded from statistical and computational analyses requiring complete datasets.

All modified Mercuri scores were assigned by a single evaluator who underwent institutionally standardized training in semi-quantitative skeletal muscle imaging assessment. Scoring was performed systematically across all imaging studies using consistent anatomical landmarks and uniform grading criteria.

### 2.4 Modeling of Impact of Protein Domain Involvement and Immunohistochemistry

Protein structural modeling was performed using ColabFold, an implementation of AlphaFold2^15^, to predict the structural impact of previously unreported CAV3 variants. Predicted structural alterations were qualitatively compared with caveolin-3 immunohistochemistry (IHC) findings obtained as part of routine diagnostic muscle pathology evaluation (1:500, Clone 26, BD Transduction Laboratories) ^16^.

### 2.5 Statistical and Computational Analysis

Principal component analysis (PCA) was conducted as a multivariate dimensionality reduction approach to evaluate sample segregation and to identify clustering patterns among muscles based on severity distribution. Hierarchical agglomerative clustering of mMS was performed using the ComplexHeatmap^17^ package (v2.14) in R^18^ to visualize patterns of muscle involvement across imaging studies.

Multivariate differences in sample grouping derived from PCA were assessed using permutational multivariate analysis of variance (PERMANOVA) with 10,000 permutations. Pseudotime scores were derived from the primary PCA axis (PC1) and used to model ordered disease progression. Monotonic ordering across progression paths was assessed using Kendall’s tau correlation. Associations between pseudotime scores and age were evaluated using simple linear regression. Linear mixed-effects modeling (LMM) was applied to evaluate the effect of severity stage on PCA-derived pseudotime scores. Pairwise multivariate comparisons between severity stages after application of pseudotime to grouping and severity were performed using Hotelling’s T² tests. Cluster separation to distinguish a predominant (typical) from a smaller atypical group was assessed using the Mann–Whitney U test (univariate, with rank-biserial correlation reported as effect size). Within-cluster stage separation was evaluated using PERMANOVA. Longitudinal progression within clusters of the typical group was assessed using Spearman rank correlation. For the atypical group, permutation testing was used to assess separation between monotonic and static muscle clusters.

Receiver operating characteristic (ROC) curve analysis^19^ was performed to evaluate the discriminative performance of representative muscles across assigned severity stages using mMS as predictor variables. Area under the curve (AUC) values were calculated to quantify discrimination. To assess monotonic progression across ordered severity stages, Spearman’s rank correlation coefficient (ρ) was calculated between mMS and stage. P-values were adjusted for multiple comparisons using false discovery rate (FDR) correction.

All analyses were performed using R version 4.4.3 within RStudio version 2023.06.1+524.

### 2.6 Data Visualization, Illustration, and Annotation

Post-analysis annotation and figure preparation were performed using Adobe Photoshop (version 23.4.1 Release), Microsoft® PowerPoint for Mac (version 16.78.3), and Microsoft® Excel for Mac (version 16.78.3).

## 3. Results

### 3.1 Demographics and Clinical Findings

Fourteen imaging study samples from eight patients were included, each assigned a unique imaging study identifier (Sample ID). Six samples were derived from pediatric patients (2.8–15 years) and eight from adults (19–61 years). Symptom duration prior to inclusion in the muscle repository ranged from 1 to 55 years. Initial clinical presentations were heterogeneous; however, two pediatric patients and one adult patient presented with fever and elevated CK levels. No patient exhibited clinical features suggestive of rippling muscle disease. Detailed clinical and genetic characteristics are summarized in Table 1.

### 3.2 Variant Classification

Five of the eight patients harbored previously unreported CAV3 variants. Identification of variants was based on available literature and database review^20–23^. The most frequently affected region was the scaffolding domain (n = 5), followed by the N-terminal cytoplasmic domain (n = 2). One patient demonstrated involvement of both the C-terminal cytoplasmic domain and the membrane-embedded domain. All of the variants were classified as pathogenic or likely pathogenic based on the American College of Medical Genetics and Genomics (ACMG) classification.

Comparison of specific mutation with immunohistochemistry findings revealed that caveolin-3 expression was absent in the muscle sample of Patient F (Sample ID 13), who carried a homozygous frameshift variant, c.87delC (p.Lys30ArgfsTer7). In contrast, Patient H (Sample ID 3) exhibited markedly reduced caveolin-3 expression, with fewer than 5% of myofibers showing faint and patchy immunoreactivity. This patient harbored two heterozygous mutations: c.436del (p.Val146Cysfs*107), a frameshift variant, and c.291C>A (p.Phe97Leu), a missense variant. Parental data were unavailable. However, upon TA cloning, the two variants were not observed in the same allele. Patient A (Sample ID 1) and Patient E (Sample ID 12), both carrying missense variants resulting in p.Ser53Arg, showed similarly diminished staining with sparse patchy positivity. Patient C (Sample IDs 4–6), harboring c.173G>C (p.Trp58Ser), exhibited diffusely weak caveolin-3 expression across examined myofibers.

Comprehensive variant interpretation, immunohistochemistry findings, and AlphaFold2-based structural modeling relative to wild-type caveolin-3 were provided in the Supplementary Data (CAV3 Pathogenicity Identification Based on ACMG Guidelines for Patients with Previously Unreported Variants).

### 3.3 Muscle Imaging Findings

#### 3.3.1 Severity Classification and Progression Modeling

The PCA of mMS-based muscle involvement profiles per Sample ID demonstrated clear spatial segregation of samples along PC1 (51.1%), forming distinct groups (Figure 1A). Samples partially clustered into discrete regions rather than forming a continuous scatter, with Sample IDs 4–6 separating from the remainder of the cohort.

**Figure 1.**
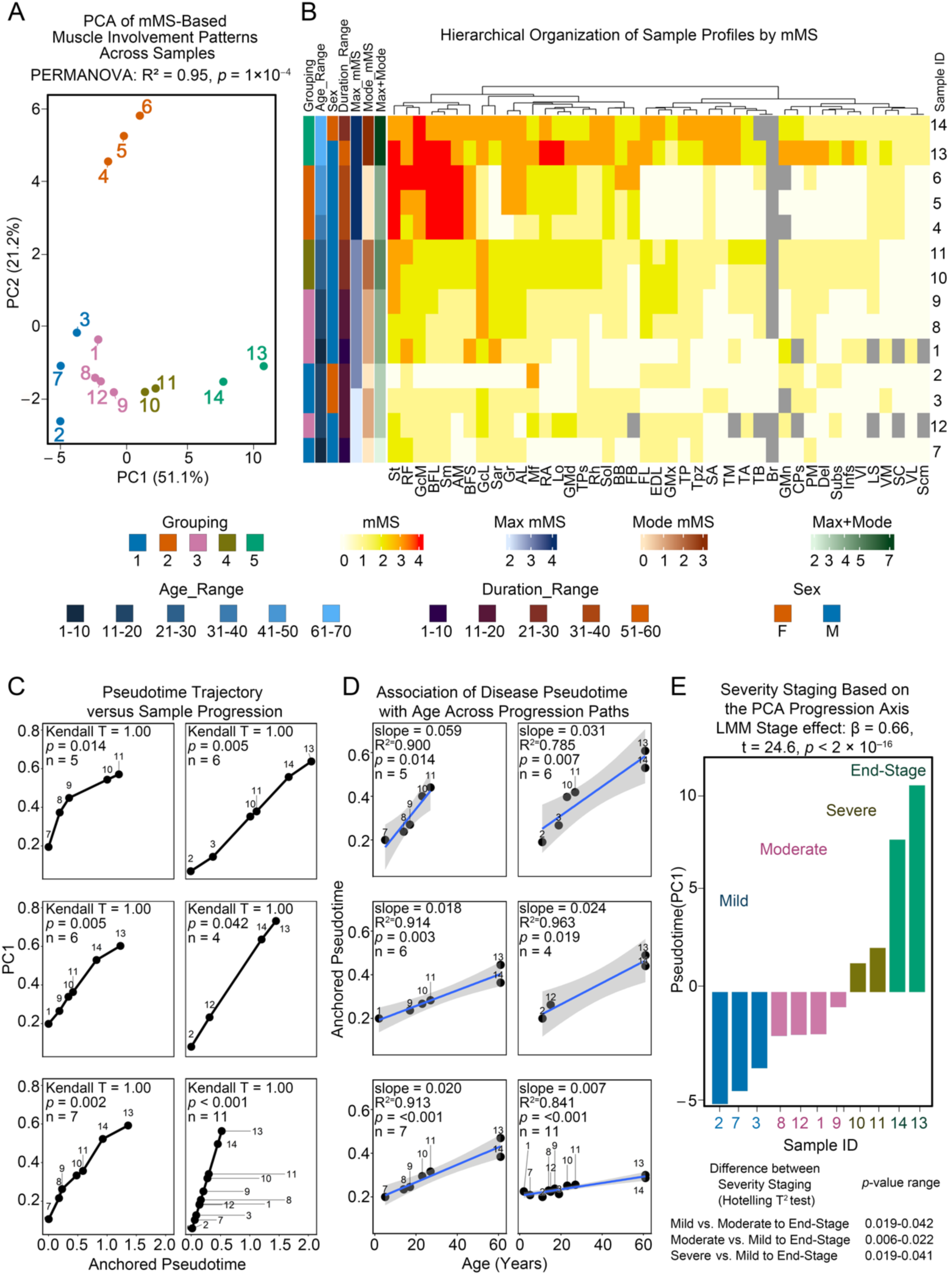
PCA-based pseudotemporal staging reveals structured progression patterns in CAV3-related myopathy based on mMS. **(A)** Principal component analysis (PCA) of mMS-based muscle involvement across samples. PC1 (51.1%) represents the primary progression axis and PC2 (21.2%) secondary variance. Samples are labeled by Sample ID and colored by grouping. PERMANOVA confirms significant separation. **(B)** Hierarchical clustering of mMS profiles (0–4) with annotations for grouping, age range, sex, disease duration, maximum mMS, Mode mMS, and combined Max+Mode index. **(C)** Concordance between PCA-derived anchored pseudotime and observed progression paths. Kendall’s τ = 1.00 across paths (*p*-values shown), indicating perfect monotonic ordering. **(D)** Association between anchored pseudotime and age within progression paths. Linear regression demonstrates significant positive relationships (R² and *p*-values indicated). **(E)** Severity staging along the PC1-derived pseudotime axis stratifies samples into Mild, Moderate, Severe, and End-stage. Linear mixed model analysis confirms a significant stage effect (β = 0.66, t = 24.6, *p* < 2 × 10⁻¹⁶), with significant pairwise Hotelling T² differences between stages.

PERMANOVA confirmed strong between-group separation (R² = 0.95, *p* = 1 × 10⁻⁴). Hierarchical clustering (Figure 1B) indicated that this segregation reflected differences in the distribution of muscle involvement rather than peak severity alone. Sample IDs 2, 3, and 7 (Group 1) showed the lowest Max mMS. Sample IDs 1, 8, 9, and 12 (Group 3) demonstrated variable Max mMS with relatively higher Mode mMS compared to Group 1. Sample IDs 10 and 11 (Group 4) shared similar Max mMS with Group 3 but exhibited higher Mode mMS. Sample IDs 13 and 14 showed the highest Max and Mode mMS (Group 5). In contrast, Sample IDs 4–6 (Group 2) reached Max mMS = 4 but maintained Mode mMS = 0, reflecting selective involvement of specific muscles (AM, St, Sm, BFL). Given these distinct patterns, samples in Groups 1, 3, 4, and 5 were designated as the typical group. These correspond to Sample IDs 1–3 and 7–14. Sample IDs 4–6 in Group 2 were classified as the atypical group to facilitate comparison between the two distribution patterns.

Pseudotime trajectory analysis excluding Sample IDs 4–6 (Figure 1C) demonstrated ordered progression consistent with Max and Mode mMS combinations (Kendall’s τ = 1.00, indicating perfect monotonic ordering within this cohort, *p* < 0.05 across paths). Anchored pseudotime showed significant positive associations with age (slope range 0.007–0.059; R² = 0.785–0.963; *p* < 0.05) (Figure 1D).

Based on these structured groupings, severity stage (Mild, Moderate, Severe, and End-stage) was assigned (Figure 1E), supported by a strong linear mixed model stage effect (β = 0.66, *p* < 2 × 10^−16^) and significant Hotelling T² comparisons (*p* < 0.05).

#### 3.3.2 Comparison of Muscle Progression for the Typical Group

Within the typical group (Mild: 2, 3, 7; Moderate: 1, 8, 9, 12; Severe: 10, 11; End-stage: 13, 14), PCA of muscle involvement demonstrated segregation of muscles into two principal clusters (Figure 2A). Mann–Whitney U testing confirmed significant separation between clusters (rank-biserial correlation = 0.24, *p* = 0.028), indicating systematic differences in patterns of involvement.

**Figure 2.**
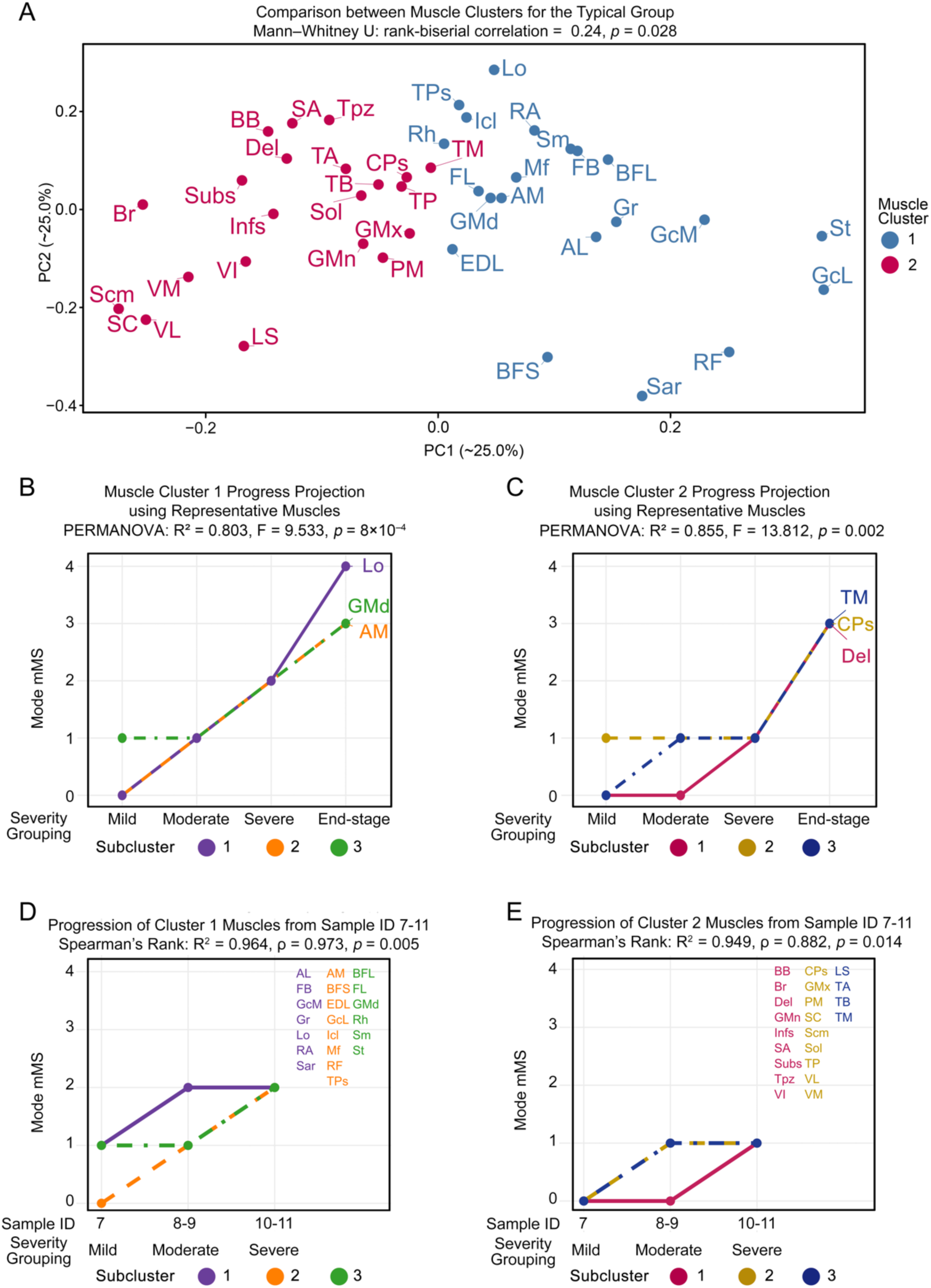
Differential progression patterns between muscle clusters in CAV3-related myopathy from the Typical Group (Sample ID 1–3, 7–14) **(A)** PCA of muscle involvement patterns within the typical group identifies two muscle clusters (Cluster 1, blue; Cluster 2, red). Mann–Whitney U testing demonstrates a significant difference between clusters (rank-biserial correlation = 0.24, *p* = 0.028). **(B–C)** Progress projection across severity stages (Mild, Moderate, Severe, End-stage) using representative muscles from each cluster. Both clusters show progressive increases in Mode mMS, with significant separation across stages by PERMANOVA. **(D–E)** Longitudinal progression of Cluster 1 and Cluster 2 muscles across samples from Sample ID 7-11 (longitudinal study of single patient). Spearman correlation confirms strong monotonic increases in mMS with advancing severity for both clusters (Cluster 1: R² = 0.964, ρ = 0.973, *p* = 0.005; Cluster 2: R² = 0.949, ρ = 0.882, *p* = 0.014).

Projection of representative muscles across severity stages (Figure 2B–C) demonstrated progressive increases in Mode mMS from Mild through End-stage. Muscles in Cluster 1 reached a peak Mode mMS of 4, whereas Cluster 2 peaked at Mode mMS 3. PERMANOVA confirmed significant stage separation within both Cluster 1 (R² = 0.803, F = 9.533, *p* = 8 × 10⁻⁴) and Cluster 2 (R² = 0.855, F = 13.812, *p* = 0.002), supporting structured stage-dependent progression.

Longitudinal evaluation across the ordered typical samples from a single patient, Sample IDs 7–11 (longitudinal study of single patient) demonstrated monotonic increases in Mode mMS within clusters (Figure 2D–E) that followed the progression trend described in Figure 2B–C. Spearman correlation confirmed strong associations between severity progression and mMS (Cluster 1: R² = 0.964, ρ = 0.973, *p* = 0.005; Cluster 2: R² = 0.949, ρ = 0.882, *p* = 0.014).

Together, these findings demonstrate that the typical group exhibits structured, stage-dependent muscle progression with reproducible cluster-specific dynamics.

#### 3.3.3 Divergent Muscle Progression Pattern in the Atypical Group

The atypical group (Sample IDs 4–6) demonstrated a distinct muscle involvement architecture compared with the typical pattern (Figure 3). PCA separated muscles into monotonic and static clusters, with significant between-cluster differentiation (Mann–Whitney U, rank-biserial correlation = −0.41, *p* = 2.7 × 10⁻⁴) (Figure 3A).

**Figure 3.**
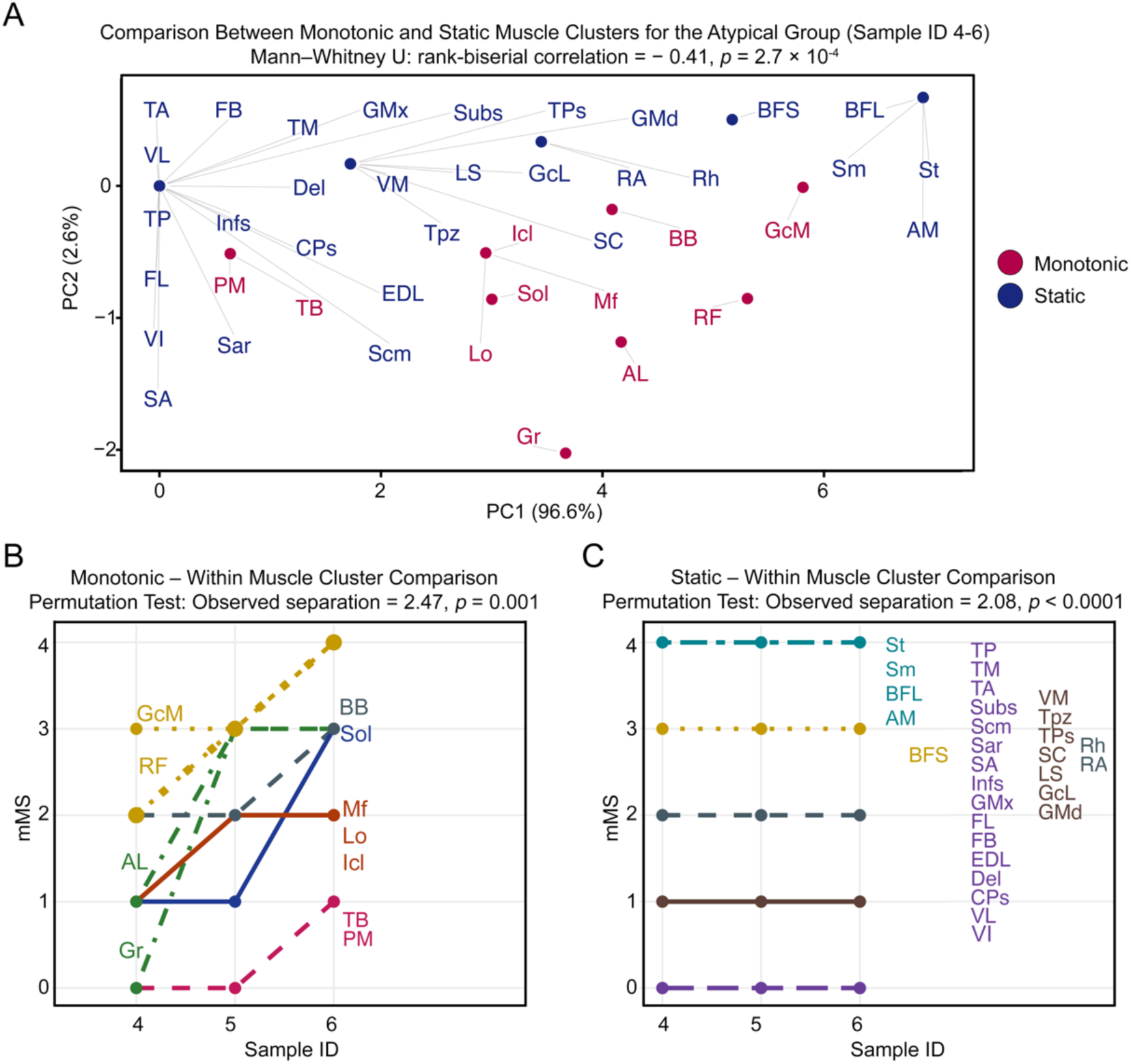
Distinct monotonic and static progression patterns in the Atypical Group (Sample ID 4–6). **(A)** PCA comparing monotonic (red) and static (blue) muscle clusters. PC1 (96.6%) captures the dominant separation between clusters. Mann–Whitney U testing demonstrates significant differentiation. **(B)** Within the monotonic group, muscle clusters show progressive increases in modified Mercuri score (mMS) across samples. Permutation testing confirms significant separation between clusters. **(C)** Within the static group, muscles demonstrate stable mMS values across samples for muscle clusters. Permutation testing similarly confirms significant separation.

Within the monotonic cluster, representative muscles showed progressive increases in mMS across stages (permutation test: observed separation = 2.47, *p* = 0.001) (Figure 3B). However, this cluster comprised fewer muscles than the static cluster. Muscles within the static cluster spanned mMS values from 0 to 4; these muscles maintained stable scores across all stages regardless of their absolute mMS value, with muscles at mMS = 0 remaining uninvolved and muscles at mMS = 3–4 remaining maximally involved throughout. A greater proportion of muscles were observed in the mMS 0–1 range (observed separation = 2.08, *p* < 0.0001) (Figure 3C).

Unlike the typical pattern, which demonstrated distributed stage-dependent progression across clusters, the atypical group was characterized by early high-grade involvement confined to specific muscles with relative preservation of more muscles across stages for the static group. This compartment-restricted architecture supports classification of Sample IDs 4–6 as a divergent progression pattern.

#### 3.3.4 Discriminative Muscle Performance and Spatial Pattern Differences

Receiver operating characteristic (ROC) analysis was performed to evaluate the discriminative performance of representative muscles across severity stages. In the typical group, multiple muscles demonstrated strong classification performance (Figure 4A). GMd and AM showed perfect discrimination (AUC = 1.00), while TM (AUC = 0.96) and Lo (AUC = 0.95) also demonstrated high accuracy. RF showed moderate discrimination (AUC = 0.86). These findings were supported by significant false discovery rate (FDR)-adjusted p-values. In contrast, the atypical group demonstrated selective discriminative capacity (Figure 4B). RF retained perfect discrimination (AUC = 1.00), whereas Lo, AM, GMd, and TM showed no significant discriminative capacity after FDR correction (Lo: AUC = 0.75, FDR > 0.05; AM, GMd, TM: AUC = 0.50), indicating absence of structured stage-dependent progression in those muscles.

**Figure 4.**
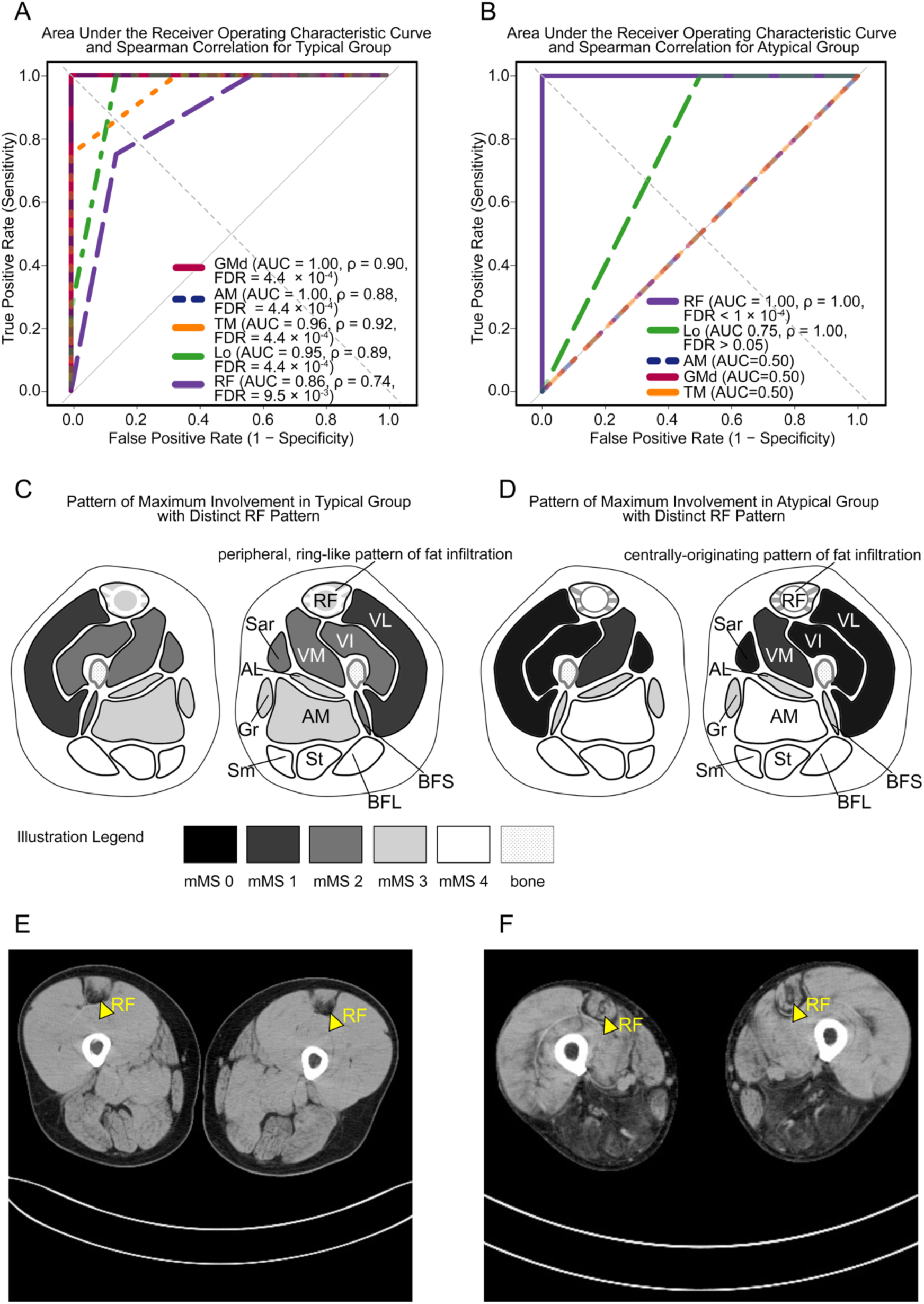
Discriminative performance and radiologic patterns of representative muscles in Typical and Atypical groups. **(A)** Receiver operating characteristic (ROC) analysis in the typical group. Representative muscles (GMd, AM, TM, Lo, RF) demonstrate high discriminative performance across severity stages, with AUC values ranging from 0.86–1.00 and significant Spearman correlations. **(B)** ROC analysis in the atypical group. RF shows perfect discrimination (AUC = 1.00) with significant Spearman correlation. Other muscles (Lo, AM, GMd, TM) show no significant discriminative capacity. **(C)** Schematic axial thigh representation illustrating the typical group pattern of maximum involvement, characterized by RF involvement with a peripheral, ring-like pattern of fat infiltration with relative central sparing. **(D)** Schematic representation of the atypical group at maximum involvement, demonstrating lesser involvement of the vasti muscles with a centrally originating pattern of fat infiltration in RF with relative peripheral preservation. **(E)** Representative axial CT image of the thigh from a typical group patient, demonstrating the peripheral, ring-like pattern of fat infiltration within the rectus femoris with relative central preservation. **(F)** Representative axial CT image of the thigh from an atypical group patient, demonstrating the centrally originating pattern of fat infiltration within the rectus femoris with relative peripheral preservation.

Schematic mapping of maximal involvement (Figure 4C–D), along with the most representative CT images (Figure 4E–F), revealed distinct spatial architectures. The most distinct pattern of fat infiltration was observed in RF, wherein the typical group exhibited a peripheral, ring-like pattern of fat infiltration with relative central preservation. In contrast, the RF of atypical group demonstrated a centrally originating pattern of fat infiltration with relative peripheral preservation. Furthermore, involvement of the vasti muscles appeared to be greater in the typical group, while AM was more involved in the atypical group.

#### 3.3.5 Anatomical Distribution of Stage-Dependent Muscle Involvement in the Typical Group

To further contextualize these findings anatomically, schematic axial illustrations demonstrated the spatial distribution of mMS–based muscle involvement across severity stages (Mild, Moderate, Severe, End-stage) in the typical group (Figure 5A–E). The end-stage group showed more muscles with higher range mMS (3–4) from the shoulder, trunk (chest wall and abdominal wall), and pelvis. Notably, only CPs showed the most evident stage-dependent increase, reaching mMS 3 at End-stage. The thigh and leg muscles showed relatively greater posterior subregion involvement.

**Figure 5.**
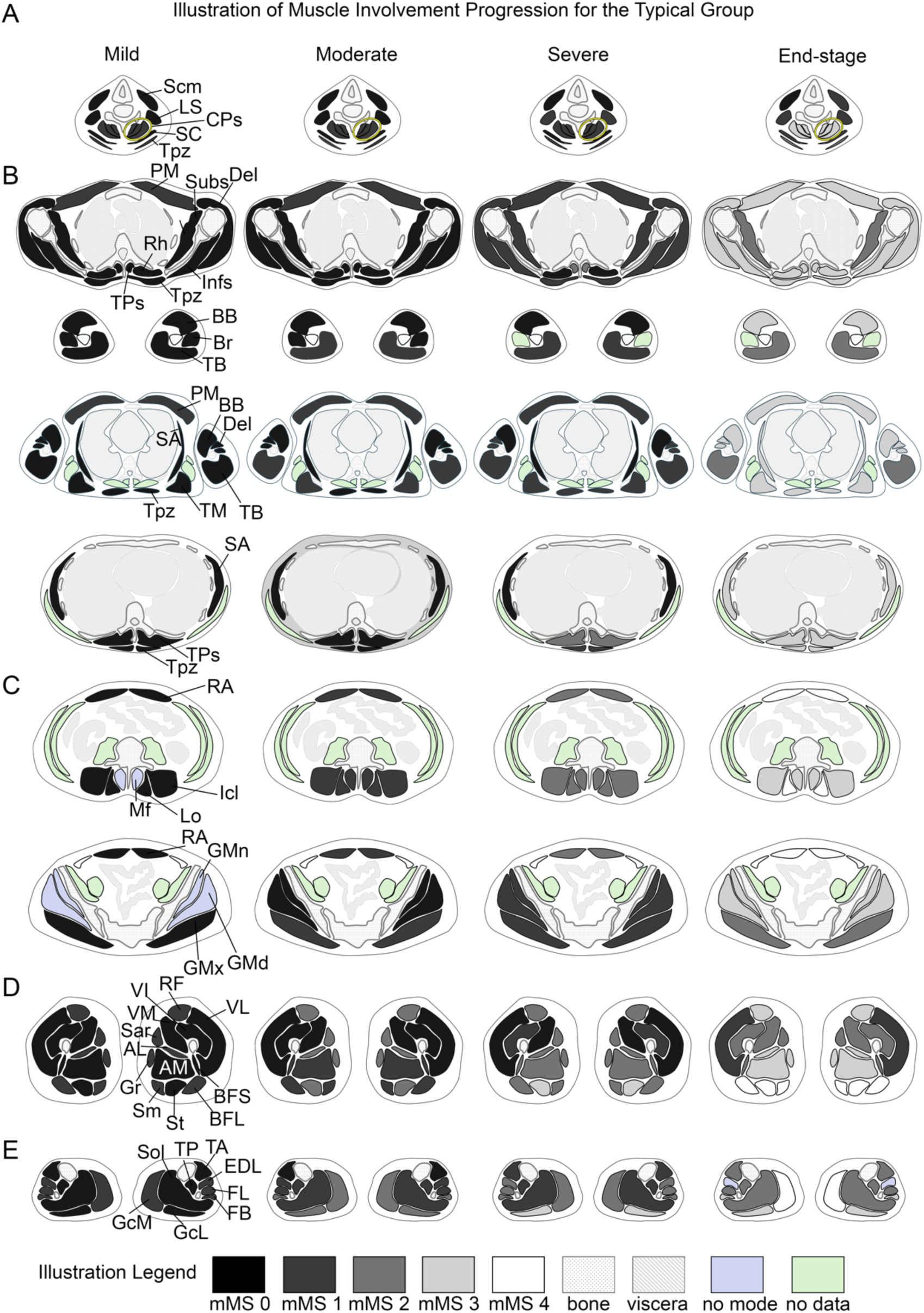
Anatomical mapping of staged muscle involvement by mMS for the Typical Group. **(A–E)** Schematic axial illustrations demonstrating the spatial distribution and progression of muscle involvement across severity stages (Mild, Moderate, Severe, End-stage). Modified Mercuri scores (mMS 0–4) are color-coded to reflect increasing fat infiltration, with additional annotations for bone, viscera, muscles without Mode mMS (“no mode”), and unavailable data (“no data”). **(A)** Neck. CPs showed mMS of 3 reached at End-stage. **(B)** Upper thoracic, shoulder, and upper arm. PM, Del, Infs, TPs, Rh, Tpz, SA, TM, and BB showed mMS of 3 reached at End-stage. **(C)** Abdominal wall and pelvic girdle. RA, Lo, Mf, IcL, GMn, and GMd reached mMS of 3 at End-stage. **(D)** Thigh. Sm, St, BFL reached mMS of 4 at End-stage. RF, Gr, AL, and AM reached mMS of 3 at End-stage. **(E)** Lower leg. GcM reached mMS of 4 at End-stage. EDL, FB, and GcL reached mMS of 3 at End-stage.

#### 3.3.6 Anatomical Distribution of Stage-Dependent Muscle Involvement in the Atypical Group

Schematic axial illustrations demonstrated the spatial distribution of mMS-based muscle involvement across severity stages (Mild, Moderate, Severe) in the atypical group (Figure 6A–E). In the neck, no apparent progression was observed across stages. More muscles in the shoulder and trunk (chest wall) demonstrated lower-range mMS (0–2). In contrast, the thigh showed higher-range mMS (3–4) in the posterior and medial compartments, along with RF.

**Figure 6.**
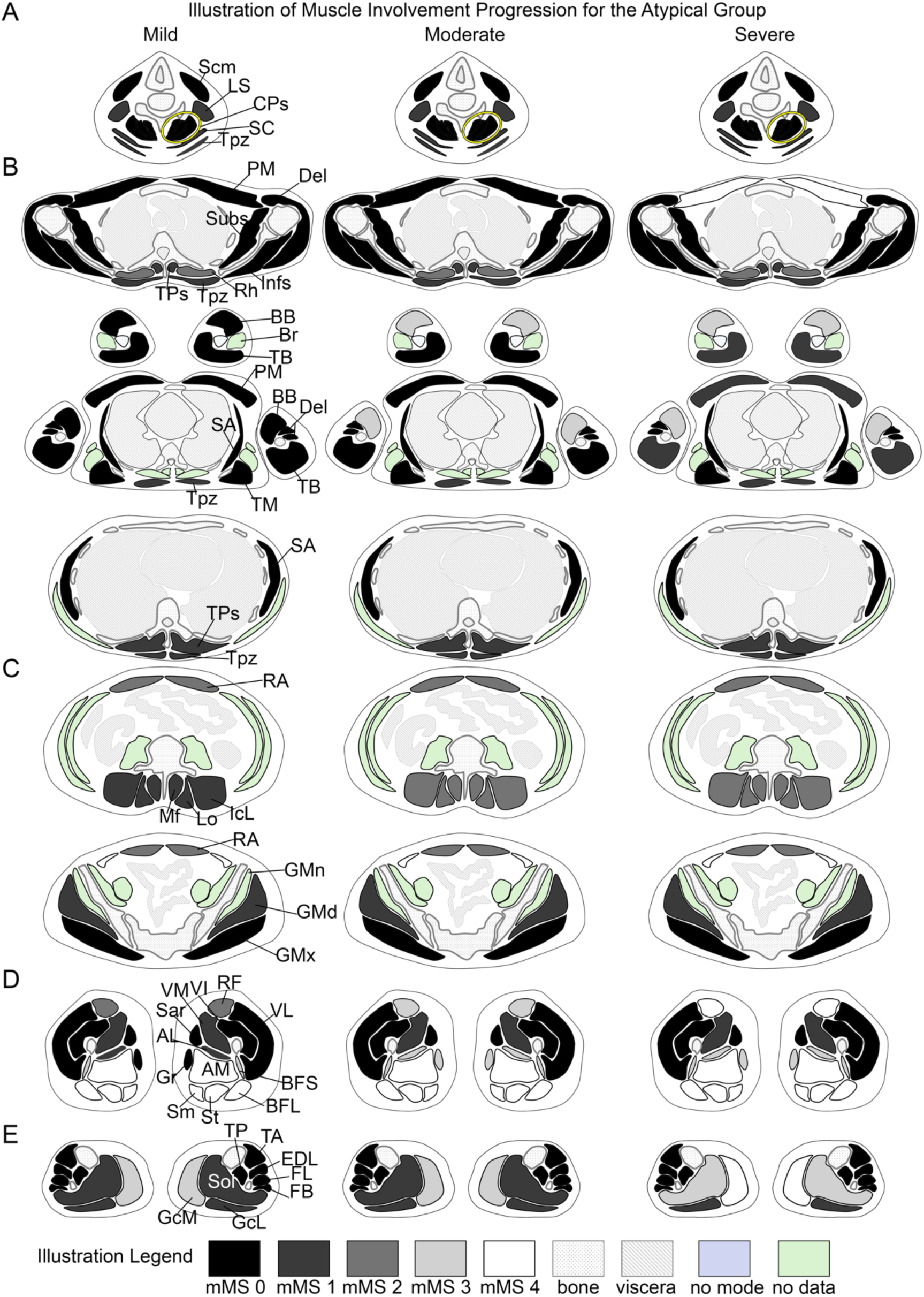
Anatomical mapping of staged muscle involvement by mMS for the Atypical Group. **(A–E)** Schematic axial illustrations demonstrating the spatial distribution and progression of muscle involvement across severity stages (Mild, Moderate, Severe). Modified Mercuri scores (mMS 0–4) are color-coded to reflect increasing fat infiltration, with additional annotations for bone, viscera, muscles without modal involvement (“no mode”), and unavailable data (“no data”). **(A)** Neck: No apparent progression for all muscles. **(B)** Upper thoracic, shoulder, and upper arm: Selective involvement of Rh (consistent mMS = 2) and early progression of BB (mMS = 3 in Moderate). **(C)** Abdominal wall and pelvic girdle. RA remains at mMS = 2 from Mild to Severe; Mf, Lo, and IcL with mMS = 2 from Moderate to Severe. Gluteal muscles consistent with mMS = 1. **(D)** Thigh. Progression of RF from mMS = 2 to 4 (Mild to Severe). AM, Sm, St, BFL with mMS = 4, and BFS with mMS = 3 throughout all stages. **(E)** Lower leg. GcM with early involvement (mMS of 3 to 4 across stages); Sol with progression to mMS = 3 at Severe stage.

## 4. Discussion

The present study demonstrates that CAV3-related myopathy without clinical evidence of rippling muscle disease may exhibit organized patterns of muscle involvement when analyzed using whole-body CT–based semi-quantitative scoring combined with multivariate modeling. Despite heterogeneity in initial clinical presentation and genetic variants, most samples followed a reproducible progression architecture, while a smaller subgroup displayed a distinct compartment-restricted pattern. By applying a systematic whole-body imaging framework, this study extended prior descriptive observation and provided a structured approach for interpreting muscle involvement across disease severity stages. The alignment of samples along an ordered trajectory, supported by multivariate analysis, suggests that the distribution of muscle degeneration in this cohort may follow a consistent progression.

In the predominant pattern observed in this cohort, muscle involvement appeared to follow an organized, selective, and partly non-uniform progression, suggesting that degeneration may occur in a pattern rather than through uniform involvement of all muscles per evaluation region. This implies differential tendency among muscle groups, in which certain muscles become involved earlier while others demonstrate comparatively delayed or attenuated progression. A smaller population of samples demonstrated a contrasting configuration in which involvement remained largely confined to posterior and medial thigh compartments. In this setting, involvement of selected muscles occurred alongside relative preservation of many other muscle groups. This may be indicative of alternative trajectories of muscle involvement. The possibility that variant-associated factors could influence distribution patterns cannot be completely excluded. However, given the limited cohort size, this observation should be interpreted cautiously. Nevertheless, maximal involvement of Sm, St, and BFL may provide an additional marker of end-stage severity that is shared across both subgroups. AM involvement pattern may further assist in distinguishing compartment-restricted from generalised disease distribution, given its greater uniformity with the posterior thigh muscles in the atypical group.

RF emerged as the most consistently informative muscle across patterns, demonstrating stage-dependent progression and distinct spatial architecture between the typical and atypical groups. Its discriminative performance across severity stages supports its utility as a representative marker of progression pattern. In the lower leg, GcM demonstrated progressive involvement across stages and may serve as a supportive marker of advancing severity alongside RF.

When compared with prior imaging studies of caveolinopathy, our findings demonstrate both concordance and extension. Ishiguro et al. described characteristic involvement of RF and posterior thigh musculature, with peripheral (“ring-like”) fat infiltration of RF reported in patients with CAV3-related myopathy, including individuals presenting with rippling muscle disease^8^. Similarly, Berling et al., in a larger multicenter cohort, reported frequent fatty involvement of Sm, St, RF, and BB, and emphasized the clinical heterogeneity of caveolinopathies^9^. In our cohort, we likewise observed the involvement of the aforementioned muscles. Furthermore, the peripheral RF pattern identified in the typical group closely parallels the radiologic architecture described by Ishiguro et al., whereas the centrally originating RF pattern observed in the atypical group represents a variation not previously characterized. Notably, none of the patients in our cohort exhibited clinical evidence of rippling muscle disease. Nevertheless, the distribution of muscle involvement closely mirrors previously reported caveolinopathy imaging patterns, raising the possibility that progressive fatty replacement in CAV3-related myopathy can occur independently of an overt rippling phenotype.

Unlike prior reports that emphasized static imaging descriptions, the present study employs a unified multivariate modeling framework to characterize and validate structured patterns of disease progression. This approach allows differentiation between a distributed limb-girdle–predominant pattern and a compartment-restricted thigh-dominant pattern, extending previous descriptive observations into a structured progression framework.

This study has several limitations. The cohort size was small and derived from a single tertiary referral center, limiting generalizability and statistical power for genotype–architecture correlations. The retrospective design precludes standardized assessment of subtle or intermittent rippling phenomena, and clinical documentation may not fully capture subclinical manifestations. Imaging analysis was restricted to semi-quantitative CT-based scoring. Given the rarity of CAV3-related myopathy, cohort sizes are inherently limited. Within this context, the multivariate framework presented here provides a structured and internally consistent model of disease progression, although validation in independent cohorts will be necessary to assess generalizability. Future studies incorporating multicenter cohorts, prospective phenotyping, and structured multivariate progression modeling may further clarify genotype–distribution relationships and determine whether progression patterns segregate independently of rippling phenotype.

## 5. Conclusions

This study demonstrates that CAV3-related myopathy without clinical evidence of rippling muscle disease exhibits structured and statistically validated patterns of muscle involvement when analyzed using whole-body CT–based semi-quantitative scoring combined with multivariate modeling. A predominant pattern was identified, characterized by a limb-girdle–predominant pattern with coordinated lower limb progression. A divergent pattern was observed in one patient, which showed a compartment-restricted thigh-dominant pattern characterized by early maximal posterior and medial thigh involvement.

The integration of semi-quantitative imaging with multivariate staging and probabilistic progression modeling provides an objective framework for characterizing disease distribution and severity compared with cross-sectional descriptive imaging analysis. These findings support the clinical utility of whole-body skeletal muscle imaging in monitoring disease progression and suggest that computational modeling approaches may facilitate more refined comparisons across caveolinopathy cohorts. Application of this framework to larger and geographically diverse populations may further clarify variability in disease distribution and progression patterns.

## Data Availability

All data produced in the present study are available upon reasonable request to the authors

## Declarations

### Ethics approval and consent to participate

All patients provided informed consent for using their samples for research after the diagnosis. This study was approved by the ethical committees of the NCNP (approval number: A2022-045, B2024-130)

### Availability of Data and Material

The data supporting the findings in this study are available from the corresponding author upon request.

### CRediT authorship contribution statement

Francia Victoria A. De Los Reyes: Investigation, Writing – review & editing, Writing – original draft, Visualization, Validation, Resources, Methodology, Formal analysis, Data curation, Conceptualization

Yoshihiko Saito: Investigation, Writing – review & editing, Writing – original draft, Visualization, Resources, Methodology, Formal analysis, Data curation

Megumu Ogawa: Investigation, Writing – review & editing, Writing – original draft, Visualization, Resources, Methodology, Formal analysis

Yasushi Oya: Investigation, Writing – review & editing, Writing – original draft, Visualization, Resources, Methodology, Formal analysis

Shinichiro Hayashi: Investigation, Writing – review & editing, Writing – original draft, Visualization, Validation, Resources, Methodology, Formal analysis, Data curation, Conceptualization

Satoru Noguchi: Investigation, Writing – review & editing, Writing – original draft, Visualization, Validation, Resources, Methodology, Formal analysis, Data curation, Conceptualization

Ichizo Nishino: Investigation, Writing – review & editing, Writing – original draft, Visualization, Validation, Supervision, Resources, Methodology, Formal analysis, Data curation, Conceptualization.

## Acknowledgments

The authors would like to thank all the patients and their attending physicians for sending muscle biopsy samples, blood, and skeletal muscle imaging data to us, and Dr. Rui Shimazaki, NCNP, for some of the template for the illustration diagram of mMS. This study was supported partly by Intramural Research Grant (5-6, 5-5) for Neurological and Psychiatric Disorders of NCNP, and Health, Labour, and Welfare Sciences Research Grants (JPMH23FC1014).

## Supplementary Data CAV3 Pathogenicity Identification Based on ACMG Guidelines for Patients with Previously Unreported Variants

**Patient A**

**Variant**: c.157A>C (p.Ser53Arg)

**Zygosity:** Heterozygous

**Interpretation**: **Likely Pathogenic** (PM2, PM1, PP2, PP3)

**Structural Impact based on AlphaFold2 Modeling (vs. Wild-type structure):** Modeling of the WT:Ser53Arg heterodimer predicted reduced inter-subunit stability compared with the wild-type homodimer, with the disruption concentrated at the scaffolding domain (aa 54–73) — the primary region mediating caveolin-3 oligomerization. The mutated residue 53 showed near-complete loss of predicted positional relationship with the opposing subunit. Neither chain appeared globally misfolded, indicating an interface-restricted rather than structural destabilization. The introduction of a bulky charged arginine at the scaffolding domain boundary is predicted to impair subunit-to-subunit contact, consistent with the partial loss of caveolin-3 expression observed on immunohistochemistry.

**Caveolin-3 Immunohistochemistry:**

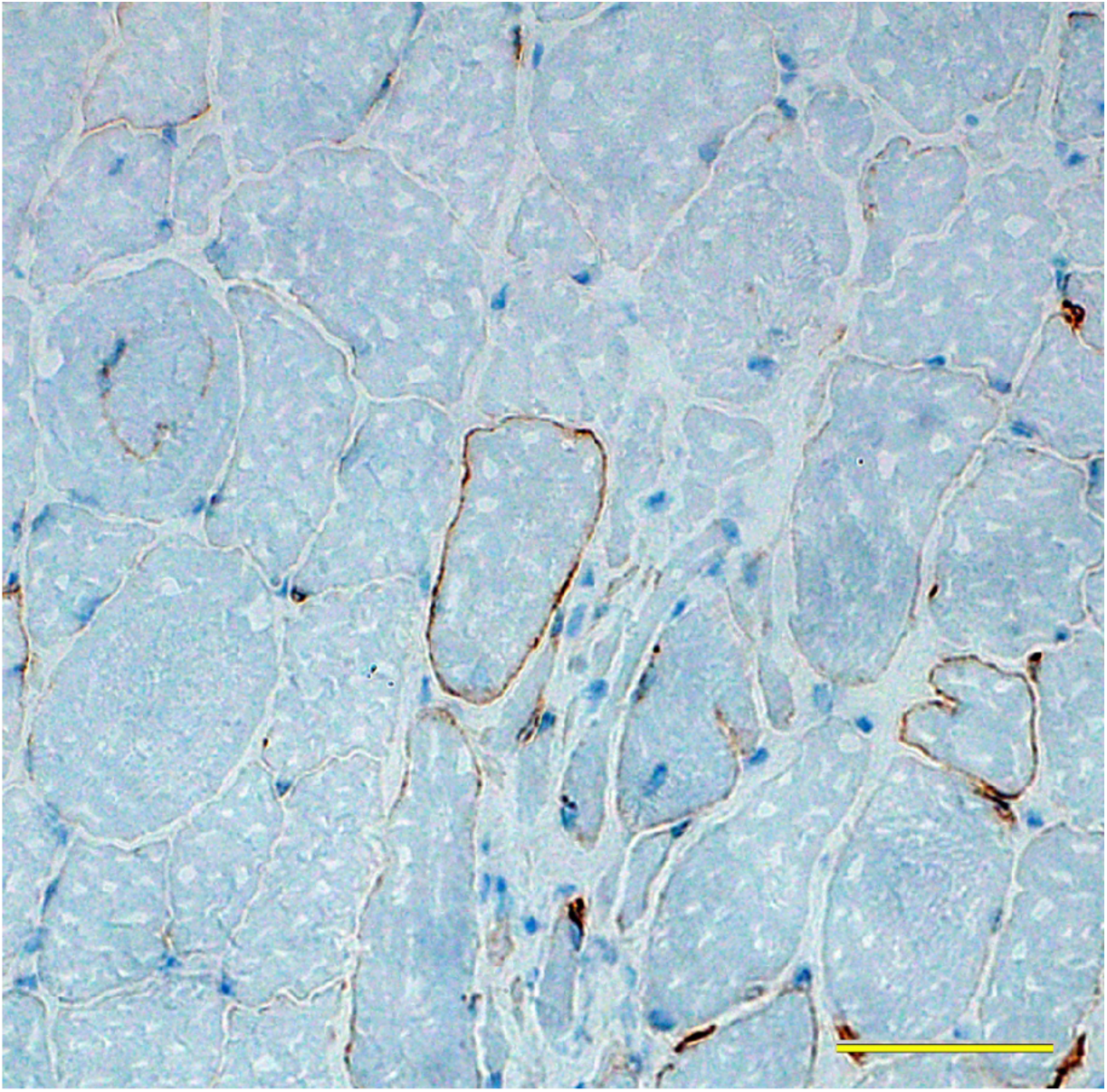

Yellow bar – 50 μm

**Patient C**

**Variant**: c.173G>C (p.Trp58Ser)

**Zygosity:** Heterozygous

**Interpretation**: **Likely Pathogenic** (PS3, PM2_supporting, PP2, PP3)

**Structural Impact based on AlphaFold2 Modeling (vs. Wild-type structure):** Modeling of the WT:Trp58Ser heterodimer predicted reduced inter-subunit stability compared with the wild-type homodimer, with the disruption concentrated at the scaffolding domain (aa 54–73) and extending into the adjacent transmembrane region. Tryptophan at position 58 contributes a key hydrophobic contact within the scaffolding domain; its substitution with the small polar serine is predicted to destabilize inter-subunit packing without disrupting the overall fold of either chain. This interface-restricted destabilization is consistent with the partial loss of caveolin-3 expression observed on immunohistochemistry.

**Caveolin-3 Immunohistochemistry:**

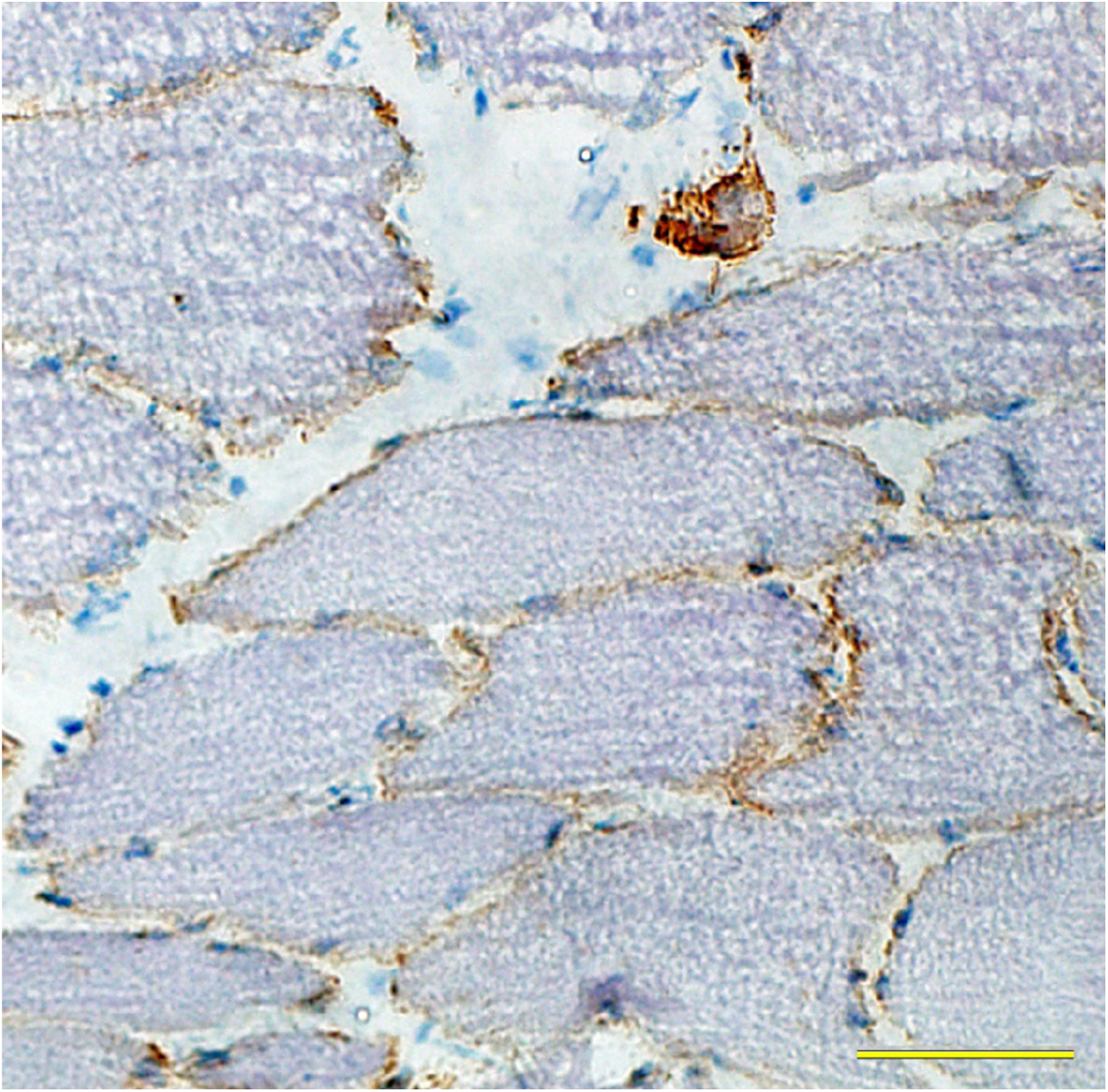

Yellow bar – 50 μm

**Patient E**

**Variant**: c.159C>A (p.Ser53Arg)

**Zygosity:** Heterozygous

**Interpretation**: **Likely Pathogenic** (PM2, PM1, PP2, PP3)

**Structural Impact based on AlphaFold2 Modeling (vs. Wild-type structure):**

Patient E harbors c.159C>A, which likewise results in the p.Ser53Arg substitution. As both variants produce an identical amino acid change, the predicted structural impact is identical to that described for Patient A above. AlphaFold2 modeling of the WT:Ser53Arg heterodimer predicted interface-restricted destabilization concentrated at the scaffolding domain (aa 54–73), consistent with the partial loss of caveolin-3 expression observed on immunohistochemistry.

**Caveolin-3 Immunohistochemistry:**

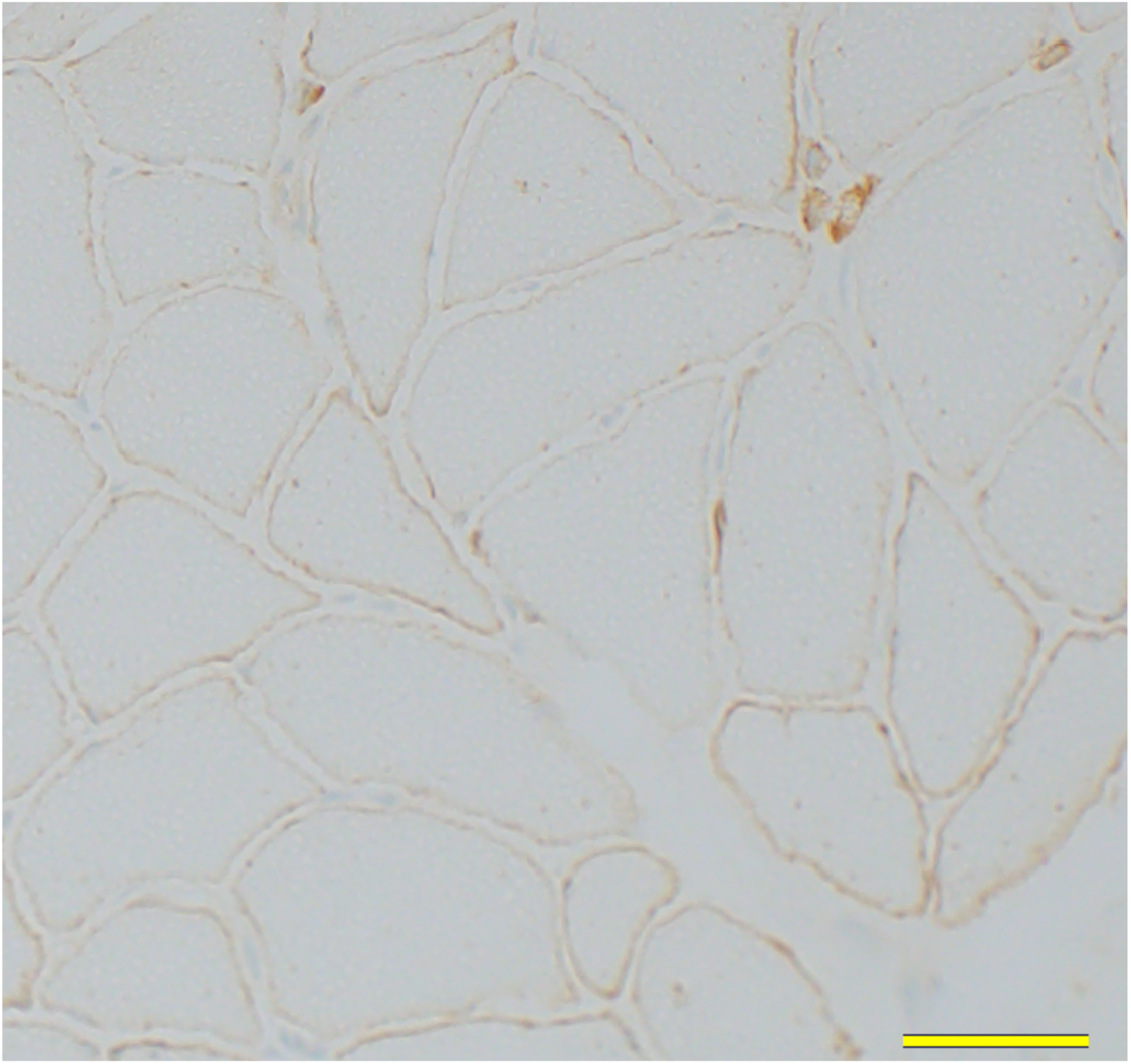

Yellow bar – 50 μm

**Patient H**

**Variant**:c.436del (p.Val146Cysfs*107) with c.291C>A (p.Phe97Leu)

**Zygosity:** Heterozygous

**Interpretation**: **Likely Pathogenic** (PVS1, PM2, PS3); **Likely Pathogenic** (PM2, PM1, PP2, PP3)

**Structural Impact based on AlphaFold2 Modeling (vs. Wild-type structure):** Modeling of the compound heterodimer (Phe97Leu:Val146Cysfs*107) predicted moderately reduced inter-subunit stability, with loss of the symmetric scaffolding domain alignment seen in the wild-type homodimer. The Phe97Leu substitution is predicted to perturb hydrophobic packing within the transmembrane region, preventing precise inter-subunit complementarity at the scaffolding domain contact surface. The Val146Cysfs*107 chain contributed a structurally disordered C-terminal extension with markedly low confidence, unlikely to support stable complex formation. Together, one allele is predicted to produce a geometrically imprecise dimer and the other a non-functional protein, providing a structural basis for the near-absent caveolin-3 expression observed on immunohistochemistry.

**Caveolin-3 Immunohistochemistry:**

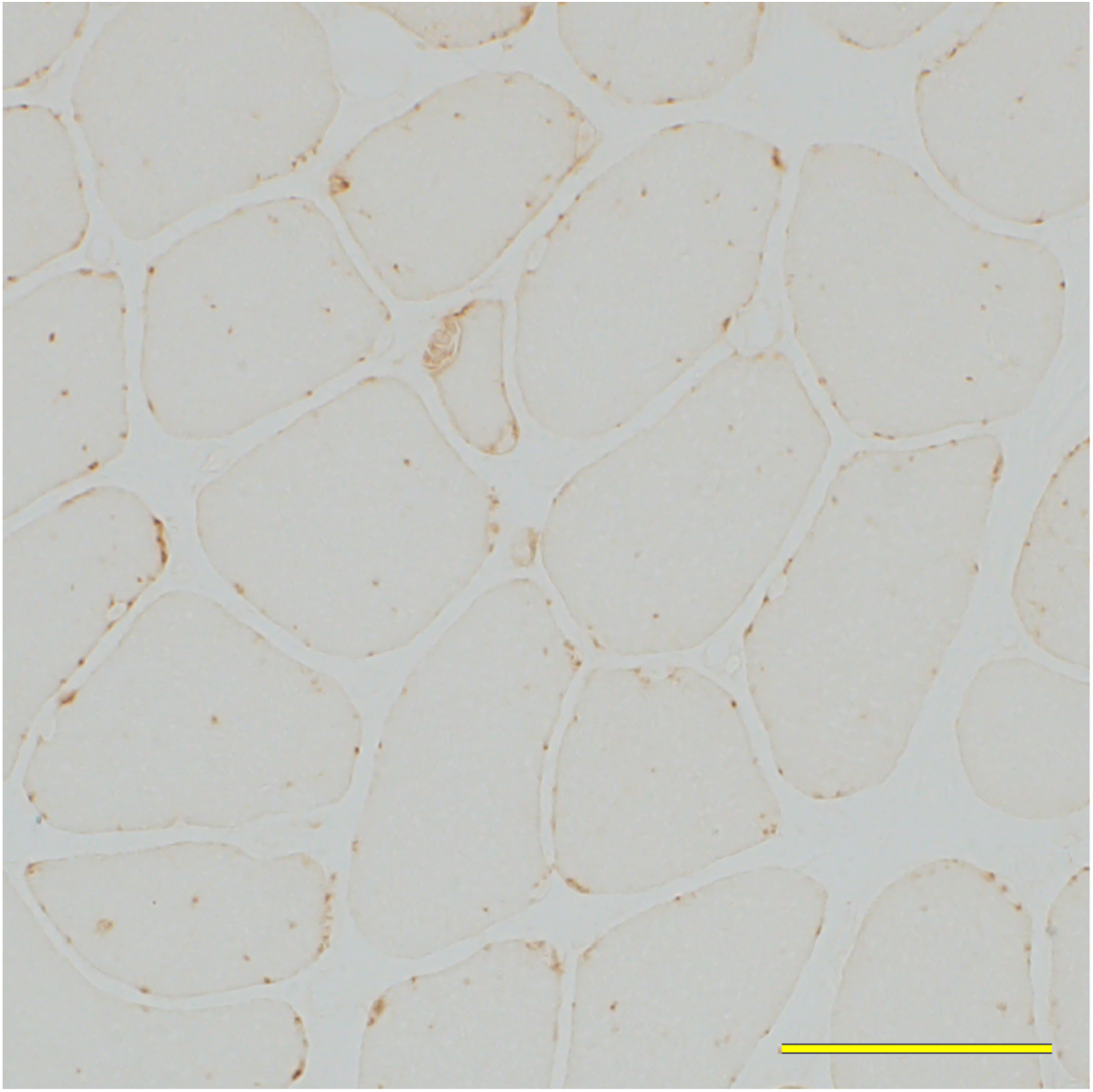

Yellow bar – 50 μm

**Patient F**

**Variant**: c.87delC (p.Lys30ArgfsTer7)

**Zygosity:** Homozygous

**Interpretation**: **Pathogenic** (PVS1, PM2, PP3)

**Structural Impact based on AlphaFold2 Modeling (vs. Wild-type structure):** Modeling of the truncated mutant homodimer predicted near-complete loss of inter-subunit organization, with overall structural confidence well below the threshold indicative of a stable protein complex. Per-residue structural confidence was uniformly low across both chains, declining progressively toward the terminal frameshifted residues. The truncation at residue 35 results in a fragment retaining only the proximal N-terminal cytoplasmic domain, with complete absence of the scaffolding domain, transmembrane hairpin, and C-terminal cytoplasmic domain. No meaningful dimer interface is predicted to form. This is consistent with the complete loss of caveolin-3 expression observed on immunohistochemistry and supports a loss-of-function mechanism through protein truncation.

**Caveolin-3 Immunohistochemistry:**

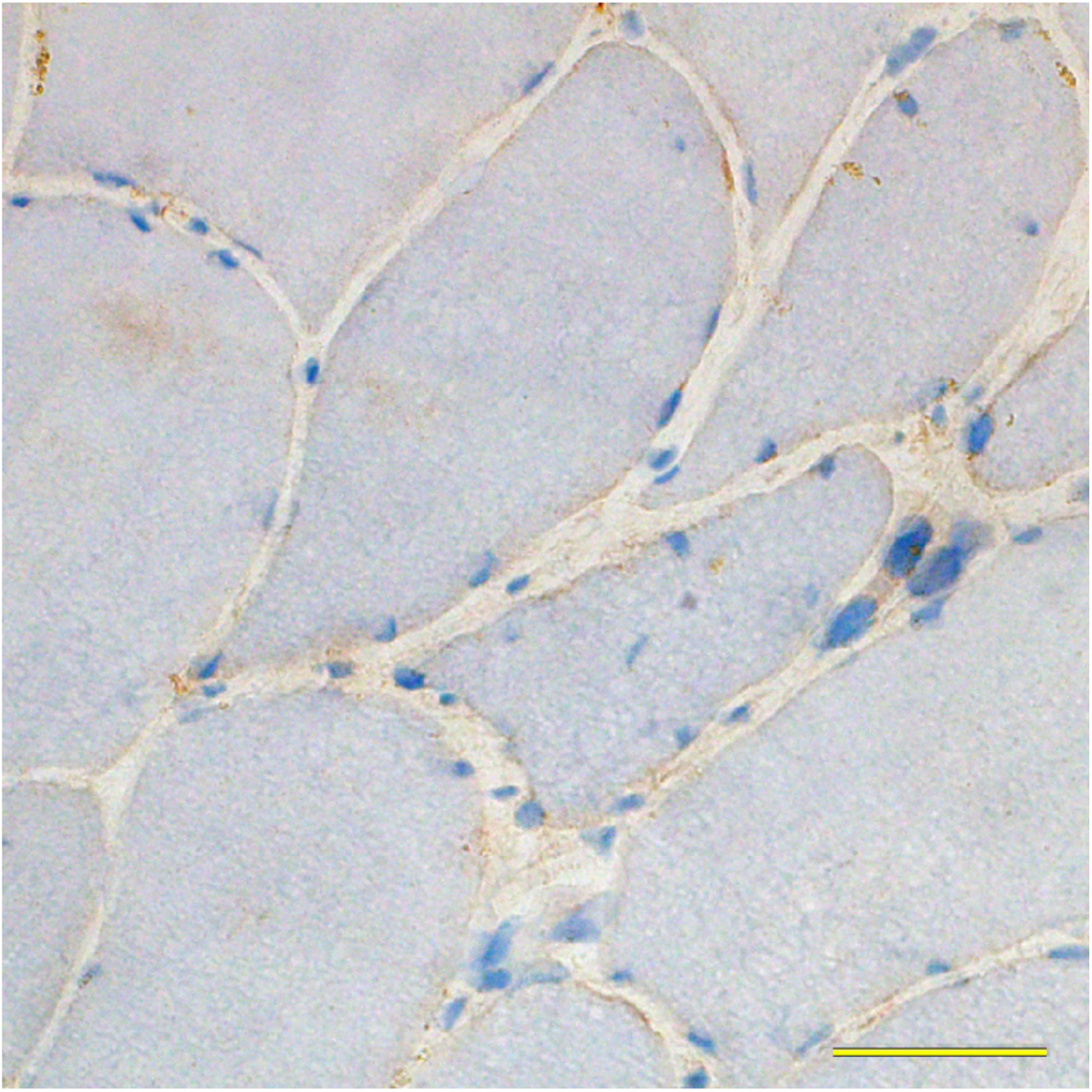

Yellow bar – 50 μm

